# The Personal Side of Clerkships: Cultural and Relational Challenges for Medical Students

**DOI:** 10.1101/2025.07.10.25331299

**Authors:** Bibi Sumera Keenoo, Naila Maherally, Anas Bin Ahmad Domun, Kashifa Rumjan

**Author notes:** **Corresponding Author:** Dr. Bibi Sumera Keenoo.

## Abstract

Clinical clerkships represent a critical phase in medical education, during which students transition from theoretical learning to immersive, hands-on practice. This stage is not only academically demanding but emotionally taxing, with students simultaneously managing professional expectations and personal relationships. While prior research has highlighted the mental health burden of medical training, few studies have examined how personal relationships influence students’ well-being and academic engagement in culturally distinct settings. This study explores the role of personal relationships, familial, romantic, and peer-based, among undergraduate medical students undergoing clinical clerkships in Mauritius, a small island nation shaped by collectivist traditions and diverse religious values.

A mixed-methods approach was adopted, combining quantitative survey data from 91 students with qualitative responses analysed thematically. Quantitative findings revealed no significant association between relationship status and demographic factors such as gender or ethnicity, but students in committed relationships reported higher levels of emotional support and relational satisfaction. Thematic analysis identified seven key themes, including cultural and religious expectations, strategic singleness, gendered emotional expression, and relationships as emotional anchors or sources of strain.

The findings underscore the socio-cultural complexity of student life during clinical training, revealing how personal relationships both buffer and exacerbate academic stress. Students’ relational decisions were strongly influenced by cultural scripts around honour, discipline, and religious morality, especially among Hindu and Muslim participants. The study advocates for culturally responsive support systems within medical institutions, including peer support networks, emotional well-being services, and relationship counselling sensitive to communal and religious dynamics. These insights contribute to a more holistic understanding of professional identity formation in medical education and call for policy reforms that embed relational awareness into student support strategies.

## INTRODUCTION

Medical education is globally recognised for its intensity, academic rigour, and the transformative demands it places upon students. At the heart of this process are clinical clerkships, structured placements that transition students from the classroom to the clinical environment. These rotations are critical for developing hands-on medical expertise, clinical judgement, and interpersonal communication skills. Yet they are also a period of heightened stress, identity formation, and emotional challenge. Clerkships not only demand cognitive excellence but also immerse students in emotionally complex scenarios involving life, death, uncertainty, and the ethical dilemmas inherent in medical practice. In this high-pressure environment, students are also expected to manage their relationships, familial, romantic, and social, alongside their professional development.

Extensive literature has documented the psychological distress often associated with this stage of training, including elevated levels of burnout, anxiety, and emotional exhaustion (Dyrbye, Thomas, and Shanafelt 2010; Ishak et al. 2013). The burden of clerkship is not experienced in isolation; it intersects profoundly with students’ personal lives. Relationships can provide essential support, acting as buffers against emotional fatigue. Equally, they may serve as sources of tension, conflict, or distraction. Navigating the expectations of loved ones while fulfilling professional obligations presents a delicate balancing act that can affect students’ academic performance, psychological well-being, and long-term personal development.

Within this global discourse, Mauritius offers a compelling and under-researched context. As a culturally diverse small island developing state (SIDS) in the Indian Ocean, Mauritius hosts a unique blend of ethnicities and religions, including Hindu, Muslim, Christian, and Creole communities. Social cohesion and family involvement are central to daily life. Among Mauritian medical students, familial expectations around academic achievement and career advancement are particularly pronounced, often taking precedence over individual desires or romantic relationships. These expectations are shaped by collectivist values, religious norms, and multi-generational living arrangements, which together create a distinctive psychosocial landscape.

For many students, particularly women, this cultural milieu can impose constraints on personal autonomy, including dating decisions, relationship disclosure, and emotional expression. In communities where academic success is deeply tied to familial pride, maintaining focus on one’s studies is often seen not only as a personal ambition but as a moral obligation. Romantic or social engagements may be viewed as distractions, or even acts of rebellion, especially for those from more conservative Hindu and Muslim backgrounds (Lovell, Lee, and Brotheridge 2015). Conversely, students from Catholic or Christian backgrounds may experience greater freedom in their interpersonal choices, reflecting subtle variations in familial and religious flexibility.

Moreover, the relatively insular nature of Mauritian society means that personal and academic spheres frequently overlap. Students often study, live, and socialise within the same closely knit communities, which can amplify feelings of scrutiny, pressure, or isolation. Decisions regarding relationships, disclosure, and self-care must often be weighed against social reputation and family honour. The result is a complex relational context in which students may struggle to maintain psychological balance, particularly during clerkships where professional and personal demands converge.

Despite the well-established emotional toll of medical education, there is a notable gap in the empirical literature exploring how personal relationships influence student experiences in culturally specific settings such as Mauritius. Much of the existing research on medical student well-being has emerged from Western, secular, and individualistic paradigms that may not reflect the realities of students in non-Western or religiously observant societies (Park, Chae, and Kim 2021). These frameworks often presuppose a degree of independence and liberal social structure that does not apply in contexts where students’ lives are deeply enmeshed with their families and communities.

As a result, key questions remain underexplored: How do Mauritian medical students experience and manage personal relationships during clinical clerkships? In what ways do cultural expectations shape these experiences, and how do different forms of relational support, or lack thereof, affect students’ ability to cope with academic stress? Without culturally sensitive insights, interventions designed to support student mental health risk being ineffective, irrelevant, or culturally inappropriate.

These questions are not only relevant locally but also resonate within the broader context of international medical education. As institutions become increasingly diverse, and as medical schools in small island states and low- to middle-income countries expand their capacity, understanding the relational and cultural dimensions of training becomes crucial. The global push for student-centred, inclusive, and context-aware education, endorsed by organisations such as the World Health Organisation, calls for research that reflects the lived realities of students from all cultural and geographical backgrounds. Mauritius, with its intricate tapestry of cultural norms and its strategic position as a regional education hub, serves as an ideal lens through which to examine these dynamics.

Furthermore, findings from Mauritius can inform policy and pedagogical strategies across similar contexts. Many small island states share socio-cultural features such as familial proximity, intergenerational responsibility, and religious influence. Understanding the interplay between personal relationships and clinical training in such settings can enhance the design of support mechanisms that foster resilience, reduce burnout, and promote well-being in culturally meaningful ways.

### Aim of the Study

This study aims to explore the influence of personal relationships on the emotional well-being and academic performance of undergraduate medical students during clinical clerkships in Mauritius.

### Objectives

1. To assess how different types of personal relationships (family, peers, romantic partners) impact medical students’ stress levels, academic engagement, and emotional well-being during clinical clerkships.
2. To examine how cultural and religious expectations in Mauritius influence students’ relationship decisions and coping strategies.
3. To identify gender- and age-related trends in how students manage the demands of clerkships in relation to personal relationships.
4. To provide evidence-based recommendations for the development of culturally appropriate support systems and interventions in Mauritian medical institutions.

## THEORETICAL FRAMEWORK AND LITERATURE REVIEW

### Situating the Study in Medical Humanities

The medical humanities explore medicine not only as a scientific practice but as a humanistic and moral encounter. Clinical clerkships represent a key phase where students are shaped not just professionally, but personally and ethically. Literature in this field has emphasised the value of narrative, reflection, and emotional awareness in medical training (Bleakley 2015; Kumagai and Lypson 2009). Clerkships bring students face-to-face with illness, death, and moral ambiguity, experiences that often intersect with their own emotional worlds, including personal relationships.

Narrative medicine, for instance, encourages students to reflect on their own emotional lives alongside those of their patients (Charon 2006). Within this context, the management of personal relationships, whether familial or romantic, becomes part of the emotional labour of medical education. Emotional repression, relational strain, or loneliness can hinder students’ ability to engage empathetically and think clearly under clinical pressure (Zaharias 2020).

Recent work has expanded on this idea. An Australian study by Harms et al. (2023) explored the burden of emotional dissonance in clinical students, finding that students with supportive personal networks were more likely to maintain professional empathy and avoid detachment. These findings support the assertion that relational well-being is integral to effective medical formation.

### Sociological Perspectives on Identity and Relational Labour

The sociology of medical training examines how institutional structures, hierarchies, and cultural norms shape the development of professional identity. Goffman’s (1959) dramaturgical theory remains particularly relevant, framing medical education as a staged performance in which students learn to present professionalism, often at the expense of authenticity. Relationships offer a “backstage” space, providing emotional release and identity reaffirmation (Rees et al. 2020).

Bourdieu’s (1986) theory of social capital is also highly pertinent. Students with greater access to relational support, be it through family, mentorship, or peer connections, are better positioned to navigate the challenges of clerkships. Recent findings by Cuff and Fields (2022) affirm this, showing that students with low perceived social capital experience higher levels of academic anxiety and identity dissonance, especially in high-pressure rotations.

The concept of emotional labour (Hochschild 1983) has also gained traction in recent literature. Medical students, especially women and students from minority backgrounds, report performing additional emotional work to align with professional expectations while concealing personal distress (Stephens et al. 2021). This labour becomes more intense in environments where personal relationships must be hidden or downplayed, as is often the case in conservative or religiously influenced settings like Mauritius.

### Cultural Anthropology: Relationality, Morality, and Identity

Informed by cultural anthropology, this study conceptualises personal relationships as culturally constructed systems of meaning rather than individual choices alone. As Geertz (1973) argued, all human behaviour, including love, friendship, and support, is mediated by cultural codes and moral expectations.

In Mauritius, collectivist traditions prioritise family harmony, social cohesion, and communal obligations. These norms influence not only who students form relationships with, but how those relationships are enacted and narrated. Religious frameworks further shape emotional disclosure and relational legitimacy. For instance, among Hindu and Muslim students, relationships outside arranged structures may be discouraged, leading to secrecy, internal conflict, or relationship breakdown (Lovell, Lee, and Brotheridge 2015).

Recent anthropological research in small island states confirms these dynamics. In a study of emotional well-being among Pacific Island medical students, Nalani and Taufa (2022) found that students often concealed emotional challenges to preserve familial honour. This cultural script of emotional suppression was linked to elevated anxiety and burnout during clerkship. Anthropologists like Kleinman (1995) have long advocated for the distinction between disease and illness experience, which calls for attention to the emotional and moral dimensions of health. Applying this to education, it becomes essential to consider not just clinical stress, but how students interpret, negotiate, and narrate their relational and emotional realities.

### Interdisciplinary Empirical Insights

Recent interdisciplinary work has strengthened the case for examining relational well-being during medical training. In their multi-country survey, Patel et al. (2021) found that medical students who perceived strong family support reported significantly lower depression scores and higher resilience. Meanwhile, a qualitative study by Singh et al. (2022) in South Asia identified relational stigma, particularly around romantic relationships, as a hidden but potent stressor for clinical students.

A 2023 review by Wearn and McKimm highlighted the urgent need to adopt culturally tailored wellness frameworks in medical education. The authors argued that globalised curricula must reflect local relational realities if they are to address mental health and burnout effectively.

### Conceptual Synthesis

Together, the insights from medical humanities, sociology, and cultural anthropology point toward a shared conclusion: medical education is not only an academic endeavour but a relational practice. Students experience clerkships not as isolated individuals, but as sons, daughters, partners, peers, and believers, embedded in cultural systems that shape their values, priorities, and emotions.

In Mauritius, these relational and cultural factors are especially salient. The weight of family expectation, the limitations imposed by religious doctrine, and the nuances of gendered experience all affect how students cope, connect, and grow during their clinical years. Ignoring these dynamics risks implementing wellness solutions that are clinically sound but culturally tone-deaf.

By grounding this study in interdisciplinary theory, we seek to capture the richness of student experience and argue for medical education policies that are both globally informed and locally attuned.

## METHODS

### Study Design and Rationale

This study adopted a mixed-methods design to explore the influence of personal relationships on the experiences of undergraduate medical students during their clinical clerkships in Mauritius. The combination of quantitative and qualitative methodologies allowed for a more holistic understanding of the complex interplay between relational dynamics and academic pressures within a culturally specific context. Quantitative methods provided baseline data on relationship status, demographics, and emotional well-being, while qualitative approaches enabled deeper insight into personal narratives, coping strategies, and cultural influences that are often overlooked in structured surveys.

This methodological approach is especially justified within the medical humanities discipline, which seeks to understand human experience in healthcare beyond clinical outcomes. By incorporating both statistical trends and individual lived experiences, this study aligns with interdisciplinary research standards that advocate for contextual sensitivity, narrative depth, and interpretive rigour.

### Participant Recruitment

Participants were recruited from the three primary medical institutions in Mauritius, all of which host clinical rotations for undergraduate students in their fourth to sixth years of study. Recruitment occurred via institutional mailing lists, classroom announcements, and student group platforms. Eligibility criteria included: current enrolment in clinical clerkships, age 18 or older, and consent to voluntary participation.

An online survey, hosted via Google Forms, was disseminated to ensure convenience and accessibility, particularly given the demanding schedules of clinical students. The survey was open for four weeks, during which follow-up reminders were circulated to enhance response rates. A total of 150 students participated, representing a broad cross-section of gender, ethnicity, religion, and relationship statuses.

### Data Collection Instruments

The survey instrument was structured in two parts:

1. *Quantitative Section*: Comprised of multiple-choice and Likert-scale questions addressing demographic data (e.g. gender, age, ethnicity, religion), relationship status (single, partnered, married, cohabiting), and indicators of emotional and academic well-being. Variables also included perceived family support, peer influence, and satisfaction with romantic partnerships.
2. *Qualitative Section*: Included open-ended prompts asking students to describe the impact of their personal relationships during clerkships, their coping strategies, and the perceived role of family, peers, and romantic partners. These responses offered contextual depth and cultural insight into how students manage the demands of medical training.

### Data Analysis

#### Quantitative Analysis

Quantitative data were exported to SPSS (Version 25) for descriptive and inferential statistical analysis. Chi-square tests were used to examine associations between relationship status and demographic variables such as gender, age, ethnicity, and years of study. One-way ANOVA was employed to assess differences in emotional support satisfaction across ethnic groups. Correlation analyses were conducted to explore the relationships between age, training duration, and relationship status.

Key statistical findings included:

- A predominant single status among students (71%), with partnered and married students representing smaller proportions.
- No significant association between relationship status and ethnicity or gender.
- Higher emotional support satisfaction among students in committed relationships.

#### Qualitative Analysis

Open-ended responses were subjected to thematic analysis following Braun and Clarke’s six-step process:

1. Familiarisation with the data
2. Initial coding
3. Theme development
4. Theme review
5. Theme definition
6. Final synthesis and reporting

Two researchers independently coded the data to ensure reliability and cross-validated emergent themes. Discrepancies in interpretation were resolved through discussion and consensus. Key themes included: familial expectations and emotional reinforcement, peer influence, relationship as distraction or support, gendered patterns in emotional expression, and religious framing of personal choices.

Themes revealed how Hindu and Muslim students often cited cultural expectations encouraging academic prioritisation and relationship avoidance, while Catholic and Christian students experienced greater flexibility in navigating relationships alongside academic duties. Gender differences were also prominent: female students more frequently relied on familial and peer emotional support, while male students tended to keep relationship matters private, emphasising career goals over personal disclosure.

### Ethical Considerations

All procedures adhered to institutional guidelines and complied with international standards for research involving human participants, in accordance with the Declaration of Helsinki.

Participation in the study was entirely voluntary, and all participants were informed of their right to withdraw at any stage without penalty. Informed consent was obtained digitally prior to the commencement of the survey.

To ensure confidentiality and data security, no personally identifiable information was collected. Participants were explicitly advised not to disclose any names or identifying details in their open-ended responses. All data were stored in encrypted, password-protected digital files, accessible only to the research team.

Given the potentially sensitive nature of questions relating to personal relationships and emotional well-being, a curated list of counselling and support resources was provided to all respondents at the end of the survey. This ensured participants had access to psychological support should any emotional discomfort arise during or after their participation.

### Patient and Public Involvement (PPI)

Following the *BMJ Medical Humanities* guidelines on patient and public involvement, this study included active input from medical students during the research design and question formulation phases. A small group of final-year students served as consultants in pilot-testing the survey, offering feedback on cultural relevance, wording sensitivity, and appropriateness of the open-ended questions.

While patients were not directly involved, the student body, as the affected public, was considered central to the research agenda. These consultations ensured the study reflected the lived experiences of the target population and informed revisions to both the survey instrument and dissemination plan.

The following PPI statement is submitted for inclusion:

> *Patient and Public Involvement Statement: Medical students were actively involved in the design and pilot testing of the survey instrument to ensure cultural relevance and appropriateness of language. Their feedback informed revisions and improved accessibility. Participants were not involved in data analysis or manuscript preparation, but results will be shared with student groups through university workshops and wellness forums.*

## RESULTS

### Participant Characteristics

A total of 91 undergraduate medical students currently undertaking clinical clerkships in Mauritius participated in this study. The gender distribution comprised 56 female respondents (61.5%) and 35 male respondents (38.5%), representing a slightly higher female representation within this cohort. Most participants were between the ages of 20 and 23, consistent with the expected age range for final-year undergraduate medical students.

In terms of ethnicity, Indo-Mauritian students constituted the majority (67%), with others identifying as Creole (15%), Sino-Mauritian (10%), and Franco-Mauritian (8%). The distribution of religious affiliation mirrored the multicultural demographics of Mauritius, with 46% identifying as Hindu, 35% as Muslim, and 19% as Christian or Catholic.

Regarding relationship status, 71% of participants reported being single, while 24% were in committed romantic relationships, and 5% were either married or cohabiting. Living arrangements varied, but most students resided with family, a reflection of the strong intergenerational and communal living practices common across Mauritian cultural groups. A small number reported living alone or with peers.

Table 1 presents the demographic characteristics of the study participants, including gender, age, ethnicity, religion, and relationship status. The diversity within this cohort provided a robust basis for interpreting how students’ personal relationships were shaped by and, in turn, interacted with socio-cultural expectations and academic demands during clinical rotations.

**Table 1.** Demographic Profile of Participants (n = 91)

| **Demographic Variable** | **Category** | **Frequency (n)** | **Percentage (%)** |
| --- | --- | --- | --- |
| **Gender** | Female | 56 | 61.5% |
|  | Male | 35 | 38.5% |
| **Age (years)** | 20–21 | 28 | 30.8% |
|  | 22–23 | 49 | 53.8% |
|  | 24+ | 14 | 15.4% |
| **Ethnicity** | Indo-Mauritian | 61 | 67.0% |
|  | Creole | 14 | 15.4% |
|  | Sino-Mauritian | 9 | 9.9% |
|  | Franco-Mauritian | 7 | 7.7% |
| **Religion** | Hindu | 42 | 46.2% |
|  | Muslim | 32 | 35.2% |
|  | Catholic / Christian | 17 | 18.6% |
| **Relationship Status** | Single | 65 | 71.4% |
|  | In a relationship | 22 | 24.2% |
|  | Married / Cohabiting | 4 | 4.4% |

### Quantitative Findings

#### Associations Between Relationship Status and Sociodemographic Factors

Chi-square tests were performed to explore the relationship between participants’ relationship status and key demographic variables. There was no statistically significant association between relationship status and gender (χ² = 3.995, *p* = 0.262), indicating that both male and female students were equally represented across relationship categories. Similarly, ethnicity did not significantly predict relationship status (χ² = 25.399, *p* = 0.384), suggesting that cultural background alone did not determine whether students pursued or maintained relationships during clerkship.

A one-way ANOVA examining the variance in emotional support satisfaction across ethnic groups also showed no significant difference (*F* = 1.152, *p* = 0.378). However, when examined qualitatively, Indo-Mauritian and Muslim students tended to describe support in terms of familial encouragement rather than romantic intimacy.

Students in committed relationships consistently reported higher satisfaction levels across several dimensions of emotional support. These included:

- Feeling understood during academic stress
- Experiencing mutual motivation
- Receiving emotional and logistical assistance
- Being able to communicate openly

In contrast, single students were more likely to report feeling isolated, especially during exams and night rotations, though some framed this solitude as beneficial to academic focus.

#### Age and Relationship Trends

The mean age of participants was 22 years (SD ±1.2), and the average age at entry into medical school was 19. A moderate positive correlation was found between age and relationship status (*r* = 0.30), indicating that older students were more likely to be in relationships, often citing stability and long-term compatibility as motivating factors. Additionally, a strong correlation was found between age and entry age (*r* = 0.61), consistent with standard educational trajectories in Mauritius.

Students in live-in relationships or marriages reported the highest levels of relational satisfaction, noting that shared living spaces facilitated daily support, reduced miscommunication, and allowed for emotional intimacy, especially during challenging periods.

The mean age of participants was 22 years, with students entering medical school at a mean age of 19. A moderate positive correlation was found between age and relationship status (*r* = .30), suggesting that older students were more likely to be in committed relationships. A stronger correlation (*r* = .61) was observed between age and entry age into medical school.

Table 2 summarises the correlations among key variables. Age was moderately positively correlated with relationship status, suggesting that older students were more likely to be in relationships. A positive correlation also emerged between relationship status and emotional support satisfaction, indicating that partnered students tended to report higher perceived support. These findings support qualitative accounts describing romantic relationships as sources of emotional stability during clinical training.

**Table 2.**
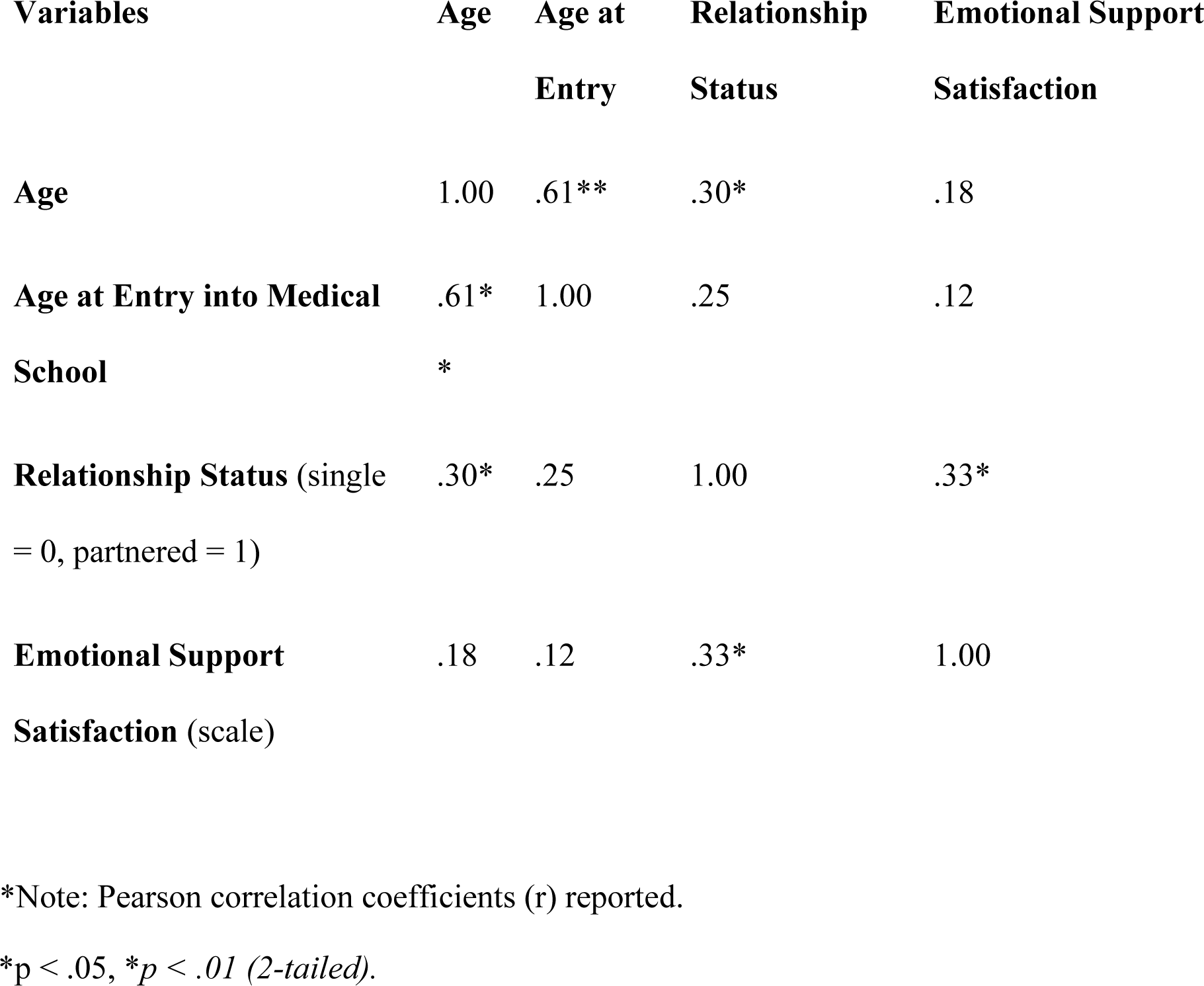
Correlation Summary Between Key Variables (n = 91)

### Qualitative Findings

Open-ended responses were analysed thematically and yielded seven primary themes and three subthemes, each capturing a unique dimension of how students experienced and managed personal relationships during their clerkships. Qualitative analysis yielded seven major themes and three cross-cutting subthemes. These are visually represented in Figure 1. The centrality of cultural and religious expectations shaped nearly all aspects of students’ emotional experiences during clinical clerkships.

**Figure 1.**
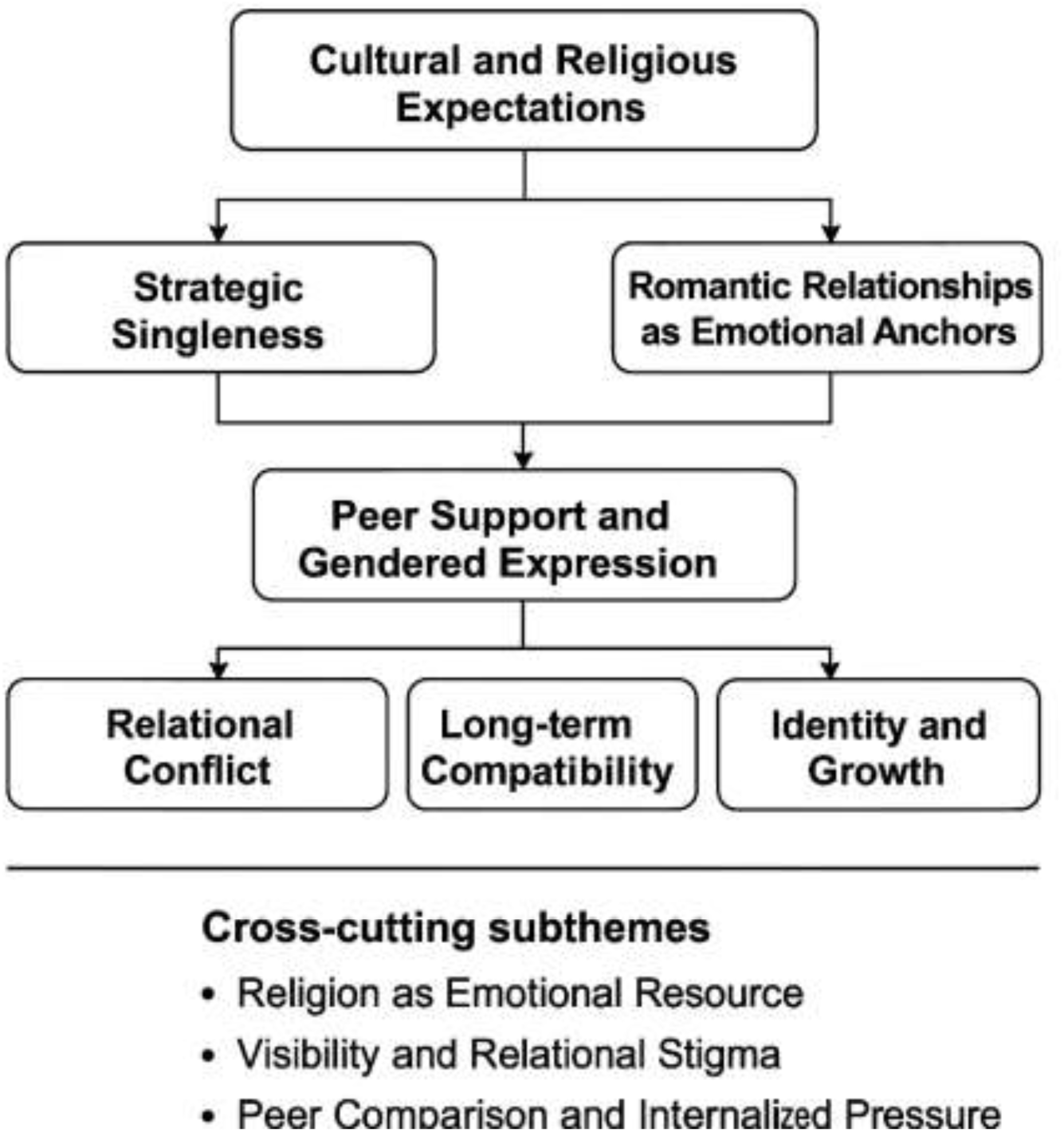
Thematic Map of Qualitative Findings on Personal Relationships During Clinical Clerkships

Figure 1 illustrates the primary themes and cross-cutting subthemes derived from students’ narratives. At the core is the influence of cultural and religious expectations, which shape students’ choices around strategic singleness and romantic relationships. These relational strategies intersect with peer support and gendered emotional expression, giving rise to downstream experiences of relational conflict, long-term compatibility, and identity development. Cross-cutting subthemes, such as religion as an emotional resource, visibility and relational stigma, and peer comparison, further nuance how students navigate emotional and academic challenges during clerkship.

#### Theme 1: Cultural and Religious Expectations

A central theme across responses, particularly among Hindu and Muslim students, was the influence of religious teachings and familial expectations. Many expressed that dating or romantic involvement was considered culturally inappropriate or discouraged unless it led to marriage. Female students, in particular, felt constrained by intergenerational norms around honour, discipline, and purity.

> *“My parents made it clear—this is the time to study, not to date.”*

> *“In my religion, dating is only acceptable when it’s serious. Casual relationships are frowned upon.”*

In contrast, Christian and Catholic students, particularly those from Creole and Franco-Mauritian backgrounds, described a greater degree of relational freedom.

> *“My parents are supportive. They’ve always said to find someone who respects me and my goals.”*

This theme illustrated the moral complexity of navigating personal autonomy within collective value systems.

#### Theme 2: Peer Support and Gendered Emotional Expression

Peer relationships emerged as a major source of emotional support, especially for female students. Friends were described as confidantes, co-survivors, and emotional mirrors, capable of offering empathy and perspective during times of clinical overwhelm.

> *“I wouldn’t have survived this year without my friends. They’re the ones who remind me I’m not alone.”*

Male students, however, tended to minimise emotional disclosure:

> *“I talk to the guys, but we don’t go into deep stuff. We just get through the day.”*

This theme reinforces the importance of gender-sensitive peer support structures and the recognition of masculine norms that inhibit emotional openness.

#### Theme 3: Strategic Singleness and Academic Discipline

Singleness was framed by many respondents as a strategic and self-disciplined choice, particularly during rotations perceived as time-consuming and emotionally demanding.

> *“Being in a relationship would be too much right now. I need all my focus for my studies.”*

This was especially common among female students, who cited both external expectations and internalised discipline as motivators. Some expressed concern about emotional instability interfering with exam performance or clinical responsibility.

#### Theme 4: Romantic Relationships as Emotional Anchors

Students in romantic partnerships often described their partners as emotional buffers, providing motivational support, encouragement, and even tangible help with academic responsibilities.

> *“He proofreads my notes, calms me down before exams, and gives me space when I need it. It’s a blessing.”*

> *“She understands my world. We’re both in the same profession, so we know what it takes.”*

These relationships often enhanced academic resilience, contradicting assumptions that romance automatically undermines focus.

#### Theme 5: Conflict and Misalignment

While many students described supportive relationships, some faced conflict and emotional drain, particularly when partners were from non-medical backgrounds or failed to appreciate the intensity of clerkship.

> *“He doesn’t get why I’m always tired or why I don’t reply for hours. It leads to fights.”*

> *“Balancing both is like juggling knives—one slip and everything crashes.”*

In such cases, relationships became another source of emotional labour, adding to rather than alleviating academic stress.

#### Theme 6: Long-Term Compatibility and Maturity

Older students in the cohort increasingly described their relationships through the lens of future compatibility, shared values, and career alignment.

> *“We’ve grown together. She’s helped shape the doctor I’m becoming.”*

> *“It’s about choosing someone who can walk the journey with you, not against you.”*

These reflections suggest that students use clerkship not only to test their professional capacity but also to assess long-term relationship viability.

#### Theme 7: Identity, Growth, and Self-Awareness

Regardless of relationship status, students described the clerkship period as one of profound personal insight and emotional growth.

> *“This year taught me more about myself than any textbook.”*

> *“Being single gave me the space to find out who I am and what I want from life—and love.”*

These accounts reinforce the idea that clerkships serve as both professional and psychosocial transitions, during which emotional patterns and relational identities are critically shaped.

### Subthemes

Three key subthemes emerged across multiple primary themes:

1. Religion as a Coping Mechanism: Students from all religious backgrounds described prayer, meditation, and spiritual reflection as essential tools for maintaining emotional balance during difficult rotations.
2. Visibility and Stigma: Students in conservative families reported hiding their relationships to avoid judgment, which created emotional strain and secrecy.
3. Comparison and Internalised Pressure: Students observing others in seemingly successful relationships expressed feelings of inadequacy or fear of missing out, particularly during moments of emotional vulnerability.

### Summary of Key Findings

- A majority of students were single, often intentionally so, viewing clerkships as incompatible with romantic distraction.
- Those in committed partnerships reported emotional, motivational, and practical benefits, particularly when relationships were rooted in mutual understanding.
- Cultural and religious norms strongly influenced relationship visibility, timing, and legitimacy, particularly among Hindu and Muslim students.
- Peer support functioned as a critical emotional outlet, though male students were less likely to utilise this resource.
- Romantic strain was more likely when partners were outside the healthcare sector and unfamiliar with the demands of clinical life.
- Clerkship was experienced not only as a professional journey but as a crucible for identity formation, emotional growth, and relational discovery.

## DISCUSSION

This study examined the influence of personal relationships on the emotional well-being and academic engagement of medical students during clinical clerkships in Mauritius. Employing a mixed-methods approach grounded in medical humanities, sociology, and cultural anthropology, the research illuminated the intricate interplay between personal relational dynamics, academic pressures, and socio-religious expectations within the unique context of a small island developing state.

The findings indicate that personal relationships, whether familial, romantic, or peer-based, are not tangential to the clinical education process; rather, they are central to how students navigate identity, stress, and performance during one of the most transformative periods of their professional formation. These relational experiences are shaped and constrained by cultural norms, gendered expectations, and institutional structures, forming a socio-emotional landscape often overlooked in traditional pedagogical frameworks.

Interpreted through an interdisciplinary lens, these findings reveal that students’ emotional and academic pathways are deeply embedded within wider socio-cultural systems. In this light, the clinical clerkship should not be viewed solely as a training phase for skill acquisition, but as a psychosocial crucible wherein autonomy, relational commitments, and evolving identity are continuously contested and negotiated.

Figure 2 illustrates this conceptual framework, highlighting the dynamic interaction between sociocultural context, institutional demands, and personal relationships in shaping students’ clerkship experiences. Medical students’ relational lives are both shaped by and responsive to prevailing cultural scripts, which in turn mediate their engagement with professional roles and emotional outcomes.

**Figure 2.**
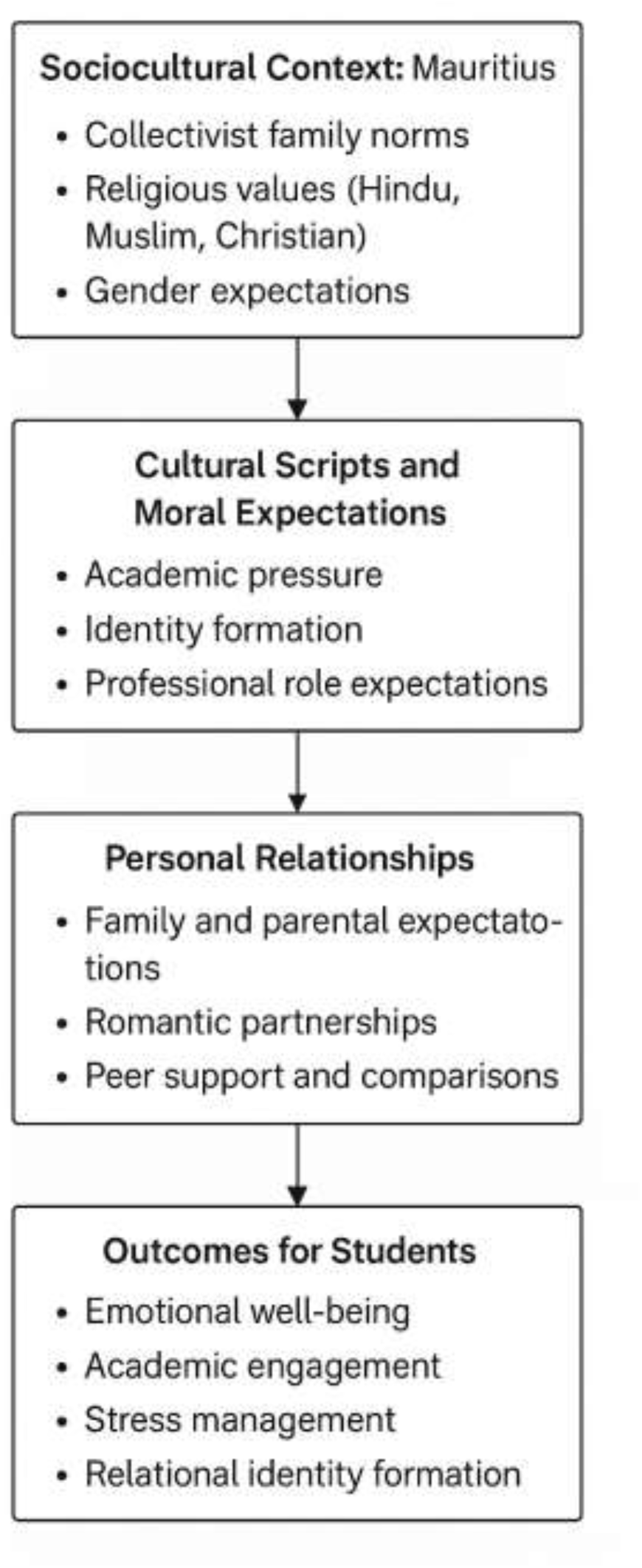
Conceptual Framework Illustrating the Influence of Personal Relationships on Medical Students During Clinical Clerkship

This framework depicts how the sociocultural context of Mauritius, including collectivist family norms, religious values, and gender expectations, shapes cultural scripts and moral expectations that students internalise during clinical training. These expectations influence the nature and dynamics of personal relationships, including familial expectations, romantic partnerships, and peer support. Together, these factors interact to affect students’ emotional well-being, academic engagement, stress management, and the development of relational identity. The model underscores the importance of culturally sensitive support systems that acknowledge the interdependence between personal, cultural, and academic domains.

### Emotional Support and Strategic Singleness

Quantitative data revealed that while a majority of students were single, those in committed relationships generally reported higher emotional support and academic satisfaction. However, many single students described their status as a conscious decision tied to academic prioritisation, reflecting a strategic form of emotional self-regulation. This suggests that the absence of romantic entanglement is not necessarily an emotional void but a calculated psychosocial adaptation to the intensity of clerkship life.

From a sociological perspective, this aligns with Goffman’s (1959) concept of role management, wherein students navigate “frontstage” academic roles and “backstage” emotional identities. For many Mauritian students, particularly women, singleness operates as a performance of discipline and conformity to familial and religious expectations. In this way, personal relationship status is not only a reflection of emotional readiness but also a negotiation of visibility and compliance.

### Cultural and Religious Frameworks

Cultural anthropology offers further insight into the normative scripts surrounding relational behaviour. In Mauritius, as in many collectivist societies, relationships are governed not only by personal choice but by a matrix of intergenerational expectations, religious values, and communal codes. Hindu and Muslim students, in particular, described pressure to defer romantic involvement until academic and financial stability were achieved, a moral logic rooted in honour, sacrifice, and filial piety.

These findings echo the work of Mauss (1925) on relational obligation and the anthropology of kinship. Relationships among Mauritian students carry not only emotional significance but also symbolic weight, functioning as vehicles for cultural preservation and moral expression. Students thus become agents navigating between traditional ideals and modern aspirations.

### Peer Support and Gendered Emotional Expression

Peer relationships emerged as essential for emotional coping, especially among female students. These findings reflect gendered socialisation patterns, wherein women are encouraged to share and process emotions collectively, while men are often expected to display stoicism and self-reliance (Stephens et al. 2021). This disparity risks leaving male students underserved by informal support networks, making them more vulnerable to internalised stress.

In the context of mental health policy, this highlights the need for gender-sensitive pastoral care strategies. Emotional well-being interventions must be designed with an awareness of how masculinity norms restrict vulnerability and inhibit help-seeking behaviours.

### Relational Conflict and Academic Identity

A minority of participants reported romantic relationships as sources of strain, particularly when partners lacked an understanding of medical training demands. These findings resonate with Hochschild’s (1983) theory of emotional labour. Students engaged in relational caregiving while managing academic stress described a “double burden” that depleted their emotional reserves and conflicted with identity formation as professionals.

This underscores the emotional dualism that clerkship students face: performing competence in clinical settings while negotiating relational roles at home. For those in unsupportive or misaligned relationships, this can result in fragmentation of identity and reduced academic engagement.

### The Role of Religious Practices in Coping

Religious rituals, such as prayer and fasting, were cited as important emotional anchors, particularly among Hindu and Muslim students. These practices offer a culturally resonant form of emotional regulation, providing spiritual structure and a sense of moral coherence amid the chaos of clerkship.

Kleinman’s (1995) distinction between the clinical and the moral in healthcare is useful here. Emotional well-being is not merely a matter of neurobiology but of meaning-making. In Mauritius, religion functions not only as a doctrine but as an affective and cognitive framework through which students interpret stress, resilience, and purpose.

### Clerkship as a Liminal Space

From a medical humanities perspective, clerkship may be viewed as a liminal period, a threshold between studenthood and professional identity. Relationships during this time are subject to constant redefinition, negotiation, and, at times, sacrifice. Whether by choice or constraint, students shape their emotional engagements in response to academic intensity and cultural legitimacy.

This interpretation finds support in recent work on professional identity formation, which emphasises emotional transitions and the importance of relational anchoring (Bleakley 2015; van den Berg et al. 2021). In this sense, students are not simply acquiring skills; they are “becoming,” restructuring their moral, emotional, and relational selves in line with the demands of the profession.

### Policy and Educational Implications

The findings have clear implications for policy and curriculum development in Mauritius and other small island states. Current wellness initiatives in medical schools often reflect Western paradigms of individual counselling, stress management, and self-directed therapy. However, these models may not translate effectively into communal, interdependent cultures where emotional well-being is interwoven with family, religion, and social roles.

To address this, institutions must develop culturally adapted wellness frameworks that validate the emotional reality of students embedded in relational networks. This includes:

- Providing confidential and culturally competent counselling services
- Facilitating peer support groups with gender-sensitive facilitation
- Offering relationship and communication workshops tailored to medical training schedules
- Engaging families and communities in student success narratives through open forums or informational materials

Additionally, academic advisors and clinical supervisors should receive training in recognising the emotional signs of relational strain, particularly among students who may be culturally constrained from seeking help overtly.

## CONCLUSION AND RECOMMENDATIONS

This study has demonstrated that personal relationships, whether present, absent, or constrained, are central to how medical students in Mauritius experience clinical clerkships. Rather than treating relationships as distractions from academic life, this research highlights their integral role in identity formation, emotional resilience, and professional development. By using an interdisciplinary lens, we have situated these experiences within the broader frameworks of medical humanities, sociology, and cultural anthropology.

Clerkship, as a high-stakes and emotionally charged stage of training, intensifies students’ reliance on interpersonal connections while simultaneously challenging their capacity to sustain them. Romantic relationships can serve as emotional anchors or sources of tension. Familial and cultural expectations can bolster resilience or impose psychological burdens. Peer support can mitigate isolation or perpetuate emotional norms. In all cases, relationships are not tangential to the medical education journey, they are embedded in it.

### Strengths and Limitations

#### Strengths

- This study is among the first to explore the influence of personal relationships on medical students in a small island developing state, offering insights that are both locally grounded and globally relevant.
- The mixed-methods approach provided both breadth and depth: quantitative data revealed patterns while qualitative data captured nuanced emotional and cultural contexts.
- The study incorporated perspectives across gender, ethnicity, religion, and age, ensuring diverse representation within the Mauritian context.

#### Limitations

- The sample was drawn from a limited number of institutions within Mauritius, which may limit generalisability to other countries or regions.
- Self-reporting via surveys introduces the possibility of social desirability bias, especially around sensitive topics such as romance, religion, and family pressure.
- Some responses were brief, limiting the richness of qualitative analysis for a small number of participants.
- As a cross-sectional study, this research cannot assess longitudinal effects of personal relationships on student outcomes such as burnout or career satisfaction.

Future studies should adopt longitudinal and comparative designs to explore these trajectories over time and across different socio-cultural environments.

### Policy Implications

The findings have important implications for both institutional policy and student support frameworks in medical schools, particularly within multicultural and communal societies.

To support student well-being in ways that reflect real-life relational contexts, medical schools should:

- Develop culturally competent, gender-sensitive wellness programmes that validate personal relationship challenges.
- Train mentors and academic staff in socio-cultural literacy, equipping them to better support students facing family or romantic pressures.
- Recognise the role of non-academic relationships in shaping performance, identity, and mental health during clinical rotations.

Additionally, embedding relational and emotional themes within medical humanities curricula may foster reflective, empathetic practice in future doctors.

### Recommendations for Educational Practice

1. Facilitate peer-led and culturally aware support spaces, allowing for open discussion of emotional challenges and relational dynamics.
2. Provide relationship management resources tailored to medical students’ schedules and stressors, e.g., seminars on communication and boundary-setting.
3. Develop anonymous digital counselling portals, particularly for students from conservative backgrounds who may be unable to seek help openly.
4. Support long-term relational planning for final-year students through optional workshops on balancing career aspirations with partnership and family life.

### Recommendations for Further Research

This study opens several avenues for future exploration:

- Comparative studies between Mauritius and other small island states or diasporic communities.
- Investigation into how LGBTQ+ students navigate relational dynamics in religiously or culturally conservative medical settings.
- Evaluation of relational stress as a predictor of academic disengagement, empathy erosion, or clinical burnout.

Moreover, a deeper exploration of religion as both a stressor and a coping resource may yield insights into the dual role of spirituality in medical training.

## Data Availability

All data produced in the present study are available upon reasonable request to the authors

## ETHICS APPROVAL STATEMENT

This study received ethical approval from the University of Mauritius Research Ethics Committee (Reference No. UOMREC/2024/112). All procedures involving human participants were conducted following institutional guidelines and the Declaration of Helsinki.

## ACKNOWLEDGEMENTS

The authors would like to thank the participating medical students for their valuable contributions, as well as the pilot-test reviewers for their insightful feedback during survey design.

